# Improving early infant diagnosis for HIV-exposed infants using unmanned aerial vehicles versus motorcycles for blood sample in Conakry, Guinea: a comparative cost-effectiveness analysis

**DOI:** 10.1101/2023.06.16.23291517

**Authors:** Maxime Inghels, Paul Mee, Oumou Hawa Diallo, Mohamed Cissé, David Nelson, Frank Tanser, Zahid Asghar, Youssouf Koita, Gabrièle Laborde-Balen, Guillaume Breton

## Abstract

**Background:** Early infant diagnosis (EID) for HIV-exposed infants is essential due to high mortality during the first months of their lives. In Conakry (Guinea) timely EID is difficult as traffic congestion prevents the rapid transport of blood samples to the central laboratory. We investigated the cost-effectiveness of transporting EID blood samples by unmanned aerial vehicles (UAV), also known as drones.

**Methods and Findings:** We conducted a cost-effectiveness comparative analysis between EID blood samples transportation by UAV compared to motorcycle using Monte Carlo simulations. Incremental cost-effectiveness ratio (ICER) per life-year gained was computed with local annual GDP per capita (US$1,194) set as the threshold. Simulation models included parameters such as consultation timing (e.g. time of arrival), motorcycle and UAV characteristics, weather and traffic conditions. Sensitivity analyses were conducted. Over a 5-year program and 778 HIV-exposed infants seen in consultation on average every year, the UAV transport strategy was able to save 49.6 life-years [90% PI: -1.3 –100.5]. The UAV strategy costs an additional of $12.4 [10.2 –14.6] per infant tested compared to the motorcycle one. With an ICER of $979 per life-year gained, the UAV transportation strategy was below the cost-effectiveness threshold. The ICER is primarily sensitive to weather-related downtime, UAV loss risk, number of HIV-infected infants and travel time saved by UAV.

**Conclusion:** The transportation of EID blood samples by UAVs in Conakry was found to be cost effective in that context. EID blood samples transportation by UAV could be a cost-effective strategy in other countries with traffic congestion and low rate of EID.

**What is already known on this topic**

- The cost-effectiveness of Unmanned aerial vehicles—UAV (drones) transportation for blood products remains limited in the Global South.
- We aimed to investigate the cost-effectiveness of urgent blood sample transportation for early HIV infant diagnosis (EID) by UAV compared to motorcycle in the region of Conakry, Guinea.

**What this study adds**

- We found that UAV transportation for EID was faster, saved more lives than motorcycles and was cost-effective (Incremental cost-effectiveness ratio: US$979 per life-year gained)
- The effectiveness of the drone strategy was primarily sensitive to weather-related downtime, UAV loss risk, number of HIV-infected infants and travel time saved by UAV.

**How this study might affect research, practice or policy**

- Our study suggests that the transportation of EID blood could be cost-effective in the Conakry region. Other low-or middle-income cities experiencing important traffic congestion with low EID could benefit from this strategy.

## Introduction

Every year, more than 150,000 children are infected with HIV and one third of these new infections occur in West and Central Africa (1). In this region, 58% of pregnant women living with HIV are receiving antiretroviral therapy (ART) and the HIV mother to child transmission rate exceeds 20% (1). Without ART initiation, new-born mortality is particularly high, especially in the second and third months of life and more than half die before their second birthday (2). Early infant HIV-diagnosis (EID) from 6 weeks of age is recommended by the World Health Organisation (WHO) in order to initiate ART before 8 weeks of age for those HIV-infected and thus significantly improving their life expectancy (3). However, only 27% HIV-exposed infants benefit from EID in West and Central Africa (1).

Because of maternal HIV antibodies persistence in the blood of infants until the age of 12-18 months, polymerase chain reaction (PCR) tests are required to provide a diagnosis. The recent development of point-of-care (POC) machines have considerably reduced PCR assay time processing and increased the number of HIV-exposed infant receiving their test result the same day as recommended by the WHO (3–5). For example, an observational study of 1793 children in Malawi showed that POC equipped sites reduced the time from sample to result from 56 days to 1 day, and allowed ARV treatment of infected children before 60 days in 91.1% of cases versus 41.9% with the standard strategy where blood sample are referred to the central laboratory (5). However, because POC sites require specific laboratory equipment and personnel, making POC cost-effective, in low HIV prevalence settings (resulting in a low test volume) is currently challenging (6). For example, in Guinea, the low prevalence of HIV (1.7%) and the decentralisation of prevention of mother-to-child HIV-transmission program to more than 300 health facilities across the country make it difficult to justify POC equipment in each site. In most African countries with low HIV prevalence, blood samples are sent to central laboratories, but the long turnaround time needed for their transportation and to send back the test result remains a major obstacle to timely EID (7,8). In addition, once the results are available, it can take up to several months for caregivers to return for the results(7,8), further delaying ART initiation among HIV-infected children and significantly increasing their risk of death. Lost to follow-up of HIV-tested children who did not received their results is also particularly high in non POC contexts and up to half of them never come back for the results (9).

To reduce the time taken to transport medical samples and supplies to inaccessible places, the use of unmanned aerial vehicles (UAV), also known as drones, has been explored in several studies in low-income countries (10–15). For example, UAVs are supplying blood bags to 20 transfusion sites located in isolated rural areas in Rwanda (15). Whilst most studies show that UAV transportation is faster but more expensive than through road (10,12,13,16), the cost-effectiveness of this strategy (i.e. consideration of life-saving benefit) remains poorly documented in low-income countries (17). In addition, UAV transportation focuses on the rural environment (10–14,17,18), while urban areas with severe traffic congestion and inadequate road networks could benefit from faster emergency delivery through UAV (16,19).

The aim of this research was to assess the cost-effectiveness of EID blood sample transportation by UAV compared to motorcycle in Conakry, the capital of Guinea, and its suburbs.

## Methods

### Study context

Conakry is an equatorial urban city in West Africa with an estimated population of 2 million (20). Conakry is located on the Camayenne peninsula which stretches out over a 40-kilometres-long strip of land 0.2 and 6.5 kilometres wide (Figure 1). The economic activities and main governmental buildings are concentrated at the tip of the peninsula accessible by a single main road which creates bottlenecks at several points in the city (21).

**Figure 1.** Map of public health centres and hospitals in Conakry and number of pregnant women testing positive for HIV in 2021, Guinea. Note 1: black dots indicate the health centres included in our analysis. Note 2: the map was generated using the Leaflet open library (https://leafletjs.com/) using OpenStreetMap (www.openstreetmap.org/) background.

At present in the Conakry region, most blood samples collected in health centres for PCR HIV-test are conveyed by road to the central laboratory which is embedded in the Donka hospital (Figure 1). The central laboratory is equipped with GeneXpert® technology for POC EID which allow to run several samples analysis at different starting time and provide a result for each test in less than two hours. To date, the main obstacle to timely EID in Conakry is the transportation of blood samples. A single van is allocated to the collection of blood samples which currently visits centres when they reach enough blood samples to collect which delay EID especially in centre with low volume of HIV-exposed children. In addition, this vehicle is often stuck in traffic jams and struggles to quickly access the various health centres in case of an emergency request.

The current plan to reduce these delays is to develop a transport system that collects blood samples for EID as soon as they are available in the health centres, sends them to the central laboratory for testing, and then returns the test result by telephone call to the health centres. The goal in this is to minimise the time of each step thus increasing the number of HIV-exposed infant receiving their result the same day and, if found to be HIV-infected, initiating ART. In the current study, we consider two type of vehicles that are adapted for the urgent transportation of blood samples: (i) a motorcycle, which is a cheaper and more flexible than a van in urban congested environments, and (ii) a UAV.

### Study design

We conducted a cost-effectiveness analysis of UAVs versus motorcycles for transporting blood samples for paediatric HIV testing in the city of Conakry. The unit of cost-efficacy was the cost for each additional year of life gained by the UAV strategy compared to the motorcycle one. We considered the implementation of both transportation strategy for a 5-year period.

We included all 33 public health centres providing postnatal care in Conakry and its suburbs in our model. An average of 1,208 pregnant women have been tested positive for HIV in these 33 facilities in 2021 (Extended data, Appendix 1). Considering the rate of HIV-exposed infants receiving a PCR test at six weeks (22), our base case assumptions model assumes an average number of 777 (min–max: 574–981) HIV-exposed infants seen in consultation at 6 weeks each year. Our study considered the different steps such as arrival of mothers in postnatal services, waiting time before consultation, time to transport the blood sample to the central laboratory, timing for test result delivery to mothers, timing of ART initiation and child survival (Appendix 2)

### Parameter and assumptions

We defined our models’ assumptions based on existing published literature, local surveys or observations and discussions with two UAV manufacturers (Table 1).

**Table 1.** Main technical assumptions for the cost-effectiveness simulation model

| Parameters | Value | [min-max] | Distribution | Sources |
| --- | --- | --- | --- | --- |
| **Healthcare** |  |  |  |  |
| Mother-to-child transmission rate at 6 weeks | 7.7% |  | Point estimate | PNLSH<sup>‡</sup> |
| HIV-exposed infant receiving PCR test at 6 weeks | 64.4% | [47.5 –81.2] | Random point estimate | (22) |
| Arrival time at the health centre | 9h00 | [7h00 – 16h30] | Beta PERT | Field survey (Appendix 3) |
| Waiting time before being seen in consultation (min) | 133.8 | SD: 66.2 | Normal with truncation of negative values | Literature review (Appendix 4) |
| Time required to perform a PCR test at the central laboratory (min) | 105 | [90 –120] | Random point estimate | Field observations with GeneXpert® |
| Work departure time of healthcare workers in postnatal services | 16h30 |  | Point estimate | Field observations |
| Mothers able to wait until healthcare workers leave | 39.0% | [15.3 –69.0] | Random point estimate | Field survey (Appendix 5) |
| Maximum waiting times for mothers (min) | 160 | [3 –600] | Beta PERT | Field survey (Appendix 5)Field survey (Appendix |
| Number of days needed to return for the test results if not collected the same day | 33 | [0 –365] | Logarithm | Literature review (Appendix 6) |
| ART initiation acceptance rates | 92.3% |  | Point estimate | (4) |
| Average age at ART initiation among children after one year | 4.7 | [1.0 –21.7] | Exponential | IeDEA data (Appendix 7) |
| **UAV** |  |  |  |  |
| UAV price | \$11,562 | | Point estimate | DeltaQuad |
| Operational horizontal speed (Km/h) | 65 |  | Point estimate | DeltaQuad |
| Operational vertical speed (Km/h) | 4.32 |  | Point estimate | DeltaQuad |
| Flight altitude required (meters) | 80 |  | Point estimate | Field observations |
| Autonomy (min) | 110 |  | Point estimate | DeltaQuad |
| Maximal flight distance (km) | 100 |  | Point estimate | DeltaQuad |
| Preparation times—e.g. UAV deployment (min) | 4 |  | Point estimate | DeltaQuad and Field observations |
| Weather inoperability | 2.6% |  | Point estimate | Meteoblue (Appendix 8) |
| UAV loss rates | 0.0% | [0.00-0.01] | Point estimate | (23), Drone Volt, DeltaQuad |
| **Motorcycle** |  |  |  |  |
| Motorcycle Price | \$3,238 | | Point estimate | CFAO motors Guinea |
| Lifespan (en Km) | 55,000 | [30,000 – 70,000] | Beta PERT | (10) |
| Breakdown rate (per 10,000km) | 0.6 |  | Point estimate | (10) |
| Minor accident rate (per 100,000km) | 1.65 |  | Point estimate | (10) |
| Major accident rate (per 100,000km) | 0.43 |  | Point estimate | (10) |
| Weather inoperability | 0.2% |  | Point estimate | Meteoblue (Appendix 8) |

CFAO: Corporation For Africa & Overseas; IeDEA; International epidemiology Databases to Evaluate AIDS; PCR: polymerase chain reaction ; SD: standard deviation; Solthis: *Solidarité Thérapeutique et Initiatives pour la Santé*; UAV : unmanned aerial vehicles <sup>‡</sup> routine data collected in Conakry, Guinea, by the National Program for HIV and hepatitis (*Programme National de Lutte contre* *Le VIH/Sida et les Hépatites*) in 2021 among 881 HIV-exposed children in public health centres and tested at 6 weeks (68 positive results)

#### Healthcare data

Healthcare data were collected through local surveys or observations and published literature. The mother’s arrival time at the health centre was collected using data on women’s attendance at two health centres (one hospital and one health centre) in Conakry (Appendix 3). Waiting times in the health centre before being seen in consultation was obtained from 5 studies conducted in Sub-Saharan Africa (Appendix 4).

Mothers were assumed to receive the results the same day if the lab test results were delivered both (i) before the average work departure time of healthcare workers in postnatal services, and (ii) before the maximum waiting time of the mothers. The maximum waiting time of mothers were obtained by conducting a local survey among 82 women (Appendix 5). When the test result was not received the same day, we created a probabilistic function to estimate the number of days before returning to get the results using data from 5 studies conducted in Sub-Saharan Africa (Appendix 6).

We assume that HIV-infected infants whose mothers do not receive the test results during the first year do not initiate ART elsewhere. For infants not having initiated ART or whose mothers were not aware of their status during the first year, we computed a probabilistic function of initiating ART at a later age (Appendix 6).

#### UAV and motorcycle parameters

We obtained data inputs from two qualified UAV manufacturers, DeltaQuad (badhoevedorp, North Holland, Netherland) and Drone Volt (Villepinte, Seine-Saint-Denis, France), both having significant experiences of unmanned aircraft system implementation in sub-Saharan Africa. UAV flight demonstrations were carried out in Conakry during the study to collect UAV real-life data and identifying specific challenges. UAV data was based on the *DeltaQuad Pro* produced by DeltaQuad (https://www.deltaquad.com/vtol-drones/map/). The *DeltaQuad Pro* is a small UAV with rotor-wing that allows vertical take-off and landing in limited space which is well adapted for urban environment. Flight routes can be planned to allow fully autonomous flights which reduce mishaps risks induced by human errors.

Motorcycle related data were collected from local dealerships such as CFAO motors (Conarky, Guinea) and local market observations. The motorcycle model chosen was the *Suzuki TF125* for its robustness and its ability to ride on rough or unpaved road.

#### Travel time

As we assumed both the UAV and the motorcycle were stationed at the central laboratory, travel times represent a return journey. We assumed that a phone call would be made from the laboratory to the health centre to inform the clinic about the test result and save additional travel times for result delivery.

Travel times by motorcycles between the central laboratory and the health centres were estimated using Google Maps data (<u>maps.google.co.uk</u>). These estimations account for travel times variability depending on months, day, time slots and travel directions (Appendix 10). After timing several travels by motorcycle between the central laboratory and the health centres, we found that the Google Maps “optimistic” estimations were closest to real-life travel times (Appendix 9). In case of breakdown or accident, we assume blood samples delivered to the laboratory the same day but related results available the following day.

Travel times by UAV were estimated using vertical and horizontal speeds as well as preparation times. We consider the UAV cruise speed to allow best speed/autonomy ratio. Given the low weight of a blood sample and its associated container (less than 50 grams), we considered its impact on the UAV speed negligible. All UAV assumptions were based on our discussion with DeltaQuad, Drone Volt and from field observations during a pilot study.

#### Weather conditions

*Meteoblue* website (www.meteoblue.com) was used to collect wind and rain data in Conakry to estimate weather operativity for UAV and motorcycle transportation (Appendix 8). *Meteoblue* is a weather forecasting website created by the University of Basel in Switzerland which can provide up to 40 years of historical hourly weather data worldwide.

We considered light rain (<2.5 mm/h) as the limit of use for the UAV and moderate rain (<7.6 mm/h) for the motorcycle. We ignored the wind strength; data shows that it has not exceeded the maximum airworthiness limit of the UAV during the last 20 years. Since we have considered a return journey, we assume that the slowing down of the UAV in case of a headwind would be compensated during the reverse trajectory of the UAV.

#### Survival

Survival curves were computed according to the infant status: (i) HIV-exposed but uninfected infant, (ii) HIV infected infant not initiating ART, (iii) HIV infected infant initiating ART. Data from the IeDEA cohort of HIV-infected children in the West Africa (24) and other existing literature were used to compute the different survival curves using double Weibull distributions (Appendix 11).

#### Costs

We devised cost functions to calculate costs from the perspective of the healthcare system (including all levels of government and donors) in 2021 US$. Euros and CFA francs costs identified have been converted to 2021 US$. All costs have been adjusted for inflation based on the year of data collection. For both UAV and motorcycle transportation strategy, the costs included purchase price, maintenance, insurance, fuel consumption, staff salaries and HIV-care related costs (for the full list, see Appendix 12). Training costs for the UAV pilot and healthcare staff were considered for UAV. We assumed that the salary for a UAV pilot in Conakry was equivalent to that for a dispatch rider employed by international NGOs in Conakry.

Residual value of the motorcycle and the UAV after the 5-year program were omitted. We assumed that the main UAV pilot will be trained at the UAV manufacturer site in the Netherlands and all additional pilots are trained by the main pilot in addition to the online course provided by the manufacturer to minimise international travel costs. We assumed that health centre staff were trained during one half-day on loading and unloading the UAV as well as security procedures. We exclude import duty and licensing costs for UAV as Guinea, like many African country, waive licensing costs for equipment and vehicles used in public health program. Because of the latter, we assumed that air navigation fees would be waived. ART cost was determined from the ART regimens prescribed and their annual cost. In Guinea, data from several paediatric HIV care specialists show that ART regimens prescribed among children are: (i) AZT+3TC+LPV/r (80%), ABC+3TC+LPV/r (10%) and AZT+3TC+EFV (10%) for children under three years of age, and (ii) ABC+3TC+EFV (100%) for those between 3 and 10 years of age. For patients over 10 years of age, ART regimens prescribed was determined using the 2016 report of the National STI/HIV/AIDS Committee on the follow-up of patient living with HIV (25) ART regimen costs were based on the price reference list published by the *Clinton Health Access Initiative* and other available materials (26,27) Non-ART care related costs (e.g. tests, consultations, fixed costs) was estimated from another study conducted in the sub-Saharan context (28).

### Analyses

The cost-effectiveness analysis was conducted using Monte-Carlo simulations. Each parameter was randomly drawn for each simulation depending on their distribution (Table 1). The unit of cost-effectiveness was the cost for each additional year of life gained by the UAV strategy compared to the motorcycle one. For each of the 10,000 simulations run for the base case scenario, the total cost and the number of survival life-year obtained were computed. Then, the differential costs between the UAV and the motorcycle transportation were divided by the survival life-year differential to obtain the incremental cost-effectiveness ratio (ICER). The threshold for cost-effectiveness per life-year gained was set at the gross domestic product per capita in Guinea, i.e., $1,194(29). A discount of 3% on both the costs and life-years gained per year was applied (30).

Sensitivity analyses were performed to identify the parameters with the greatest influence on the ICER.

Analyses were conducted using the software R version 4.0.2. (R Core Team. Vienna, Austria).

### Patient and public involvement

Patients or the public were not involved in the design, recruitment, conduct or dissemination of this study.

## Results

### Travel times

We assumed that one UAV and one motorcycle were deployed as round-trips time per day totalised on average 106 minutes [72-144] and 213 minutes [142-292] for the UAV and motorcycle respectively—which represents an average of 3 missions a day. UAV transport saved an average of 35.7 minutes in travel time and up to an average of 108.1 minutes for the most remote centres, over 50 kilometres from the laboratory (Appendix 10). No time was gained by the UAV for the two centres located within 3 kilometres range.

### Health outcomes and survival

A higher proportion test results were delivered to mothers the same day with the UAV strategy (53.0% [90% prediction interval: 34.8 –71.1] vs 47.4% [29.5 –65.4] for the motorcycle strategy). After considering the probabilities to return for the results among mothers not receiving the results (see methods, Appendix 6 and 7), an average reduction of 5.8 days for receiving the results was found for the UAV strategy (48.0 days [30.8 –65.2] vs 53.8 days [36.3 –71.4] for motorcycle). These results correspond to a reduction of 8.3 days for ART initiation (101.0 days [46.6 –155.5] vs 109.3 [52.7 –165.8] for motorcycle). Over the 5-year period program, the UAV transport strategy was able to save an additional 49.6 life-years [-1.3 –100.5] compared to the motorcycle strategy.

### Cost

Excluding treatment and HIV care costs, median investment costs were $95,141 [95,106 –95,175] and $54,550 [Interquartile range: 53,282 –56,111] for the UAV and motorcycle strategy respectively. Transport costs per blood sample were $24.6 [23.5 –25.8] by UAV and $17.3 [16.9 –17.8] by motorcycle. When considering all costs (including ART and care costs), the UAV strategy costs an additional of $12.4 [10.2 –14.6] per infant tested compared to the motorcycle strategy.

### Cost-effectiveness analysis

With the base case assumptions, the overall incremental cost-effectiveness ratio found was $979 per life-year saved which was just below the threshold we used for cost-effectiveness—i.e. $1,194 (Figure 2). The cost-effectiveness of the UAV transport strategy was superior in 64.8% of the 10,000 simulations performed. In 31.2% of simulations, the UAV transport strategy was effective but too expensive and in 4.0% situation it was less effective.

**Figure 2:** Figure 2. Cost-effectiveness plane of blood sample transportation by UAV versus motorcycle. Note: Each blue dot represents a simulation. Error bars show 90% prediction intervals.

### Sensitivity analyses

Sensitivity analyses of the main parameters showed that the cost-effectiveness of the UAV versus the motorcycle was highly dependent on four factors: (i) weather inoperability, (ii) the capacity of the UAV to be faster than the motorcycle, (iii) UAV loss risk, (iv) number of children infected (i.e. mother-child transmission rate), and (v) UAV purchase price (Figure 3). The UAV strategy stayed cost effective in the following situations: weather inoperability of 5% or less, (ii) minimum average transportation time saved by UAV of 27 minutes, (iii) UAV losses below 1 every 4,000 flight-hours, (iv) mother to child transmission equal to 5.8% or more and (v) UAV price below $15,483.

**Figure 3:** Sensitivity analysis of key parameters.

^1^Including only centres at a certain distance mechanically reduces the number of children exposed to HIV as fewer centres are included.

Note: the cost-effectiveness threshold is determined when the cost per life-year gained is equivalent to the GDP per capita (i.e., $1,194).

To allow generalisation of our result to other contexts, we ran a series of different scenarios that all consider a similar number of HIV-exposed infants seen every year in each health centre (Figure 4). Higher weather inoperability can be balanced with higher time saved by UAV transportation if a minimum of HIV-infected infant benefit from the strategy. UAV price reduction or increase have less effect on the cost-effectiveness of the UAV strategy in case of high number of HIV-infected infant benefiting from the strategy.

**Figure 4:** Cost-effectiveness of blood samples transportation by UAV depending on main parameters Note 1: Results obtained by modelling the overall cost and the number of life-years gained by the average time saved using linear regressions. Note 2: For comparison, in Conakry region, an estimated 777 HIV-exposed infants (with 60 HIV-infected) were seen in postnatal consultations in 2021, 2.6% of UAV inoperability due to bad weather, and the average time saved per UAV was estimated to be about 35.7 minutes.

## Discussion

Our results showed that transporting blood samples for paediatric HIV testing by UAV in the Conakry region was cost-effective when compared to transportation by motorcycle. The UAV transport strategy has the potential to be cost-effective in other contexts, provided there is sufficient time saved in transportation, enough HIV-infected infants benefit from the UAV transportation, and weather conditions do not result in too many downtimes.

This is one of the first study to investigate the cost-effectiveness of UAV transportation in a Global South country. UAV systems involved for medical purposes are quickly expanding in Africa (31), and mostly focus on delivering health goods (e.g., blood products, vaccines) in remote or isolated area. Our study confirms existing economic evaluations showing that UAV transportation systems are significantly more expensive compared to traditional modes of transportation(10,13,16). Nevertheless, our study is one of the first to demonstrate the possibility of UAV transportation systems to save lives while remaining a cost-effective strategy in a low-income context. Our results support the implementation of UAV transportation systems, although our analysis focused on blood sample transportation for paediatric HIV testing only. Therefore, the cost-effectiveness of UAV transportation for other blood products should be evaluated.

Although our analysis focuses on the specific case of the Conakry region, evidence from our study suggests that UAV transportation of blood samples for paediatric HIV testing is a strategy that could be implemented in other relatable contexts. First, our base case assumptions considered a mother to child transmission at six weeks of 7.7% where this rate is higher in many countries, in particular in the Sub-Saharan region (32,33). As the cost-effectiveness of the UAV strategy was particularly sensitive to the number HIV-infected infants, this suggests that this strategy may be considered in similar Global South contexts with high rates of mother-to-child HIV transmission. Second, the cost-effectiveness threshold defined in our analysis is very low because Guinea is one of the world’s poorest countries, which suggests that the UAV strategy could be cost-effective in a similar context in other low-or middle-income countries. Third, on average the UAV in our analysis flew on average for 106 minutes per day due to the limited range of the UAV but also to the localisation of the central laboratory, where the UAV was based, which does not take advantage of the full area the UAV operation radius can cover. A similar study in a setting with a more centralised laboratory or an inland rather than coastal location could increase the number of health centres reached by the UAV and thus increase the number of HIV-exposed infants benefiting from the intervention. Fourth, the UAV market and technology is not yet mature enough which entails quick evolutions in prices drop and UAV technology development which are likely to make the UAV more affordable and efficient (34). Thus, it is expected that UAV will become a more affordable and effective solution for the transport of urgent blood products in the foreseeable future.

The UAV strategy reduced by 8.3 days the mean time before ART initiation among infant leading to 49.6 life-years saved. Our results showed that UAV cost-effectiveness could be increased if a greater number of HIV-exposed children benefitted from this strategy. Such improvement would be possible by increasing the currently low postnatal consultation rate (64.4%) or by including a higher number of health centres in Conakry enabled by a higher operational range of the UAV. The current development of hydrogen batteries has recently made it possible to extend the flight times before recharge to more than 12 hours for a UAV model similar to the one used in our simulations which suggested that micro UAV will soon be able to reach more distant centres (35). In addition, considering the high down time in our simulations (on average there was only 106 minutes of flight per day) UAV could be used for other health related emergencies such as blood transportation for transfusion in the case of obstetric haemorrhage which is the leading cause of maternal death in sub-Saharan Africa (36).

The cost-effectiveness of the UAV strategy was highly dependent on weather conditions. In the city of Conakry, these conditions seem rather favourable since only 2.6% of the UAV trips were cancelled due to bad weather, well below the 10% threshold assumed by another study in a nearby country (10). It is possible that this difference is due to specific local weather contexts, the UAV’s longer distance travel (increasing its exposure to areas with frequent tropical rainfall), or the characteristics of the deployed UAV. Nevertheless, the consideration of weather conditions remains an important factor in the viability of a UAV transport strategy that is often omitted in other studies (11–13,16).

Our study fills several gaps within the existing literature on UAV in low-income countries. First, it documents the potential of UAV use in congested urban environments which is a persistent issue in developing countries (19,21). Like other studies, we found that use of a UAV significantly decreases travel times in congested traffic environments (11,16), which supports the UAV as a relevant solution for emergency transport in these contexts. Second, to the best of our knowledge, our analysis is one the firsts to include lifesaving cost benefits of UAV transportation in a low-income country (10,12,13,16). To date, we find a unique study that have investigated the lifesaving cost benefits of using of UAV transportation in the Global South. This study has investigated the use of UAV for improving tuberculosis screening in remote region of Madagascar (17). However, their analysis has several limitations, such as the omission of important factors such as weather conditions, the strong assumptions in their model regarding the effect of the drone on the likelihood of being tested for TB, and the absence of a description of the comparative transport in the standard of care. In Germany, when considering the cost of each life-year saved, a study conducted found that UAV-based delivery of defibrillation for out-of-hospital cardiac arrests was cost-effective (37). Thus, our results add to the literature another piece of evidence that UAV can be cost-effective for urgent medical transport, even in a resource-constrained environment.

Our analysis has some limitations. Data not available for the city of Conakry were assumed from other studies conducted in the Sub-Saharan context and may not represent the local context. We neglect the risk of crashing or losing the UAV in our base case assumptions, although we did conduct sensitivity analysis on that parameter. The risk of loss or accident for “small” UAVs remains poorly documented overall(10). Military UAV data suggested an accident risk of less than 5/100,000 flight hours with human error explaining the majority of these accidents (23). In all our simulations, the use of the UAV never exceeds 3,000 flight hours over the 5 year program. In addition, many UAV models, including the one used in our simulation, has autopilot systems allowing them to perform entire missions autonomously without the need for manual remote control which would limit human errors. In addition, several improvements are now available to limit flight accidents such as the equipment of sensors to avoid unexpected obstacles during the flight (38).

As our results rely mainly on sub-Saharan African data, these results could be generalised to other African settings with both high delay HIV paediatric results and traffic congestion— two salient issues in many countries in West and central Africa (1,19). EID blood sample transportation could be considered for major West African urban cities such as Abidjan or Lagos—the higher GDP per capita of their related countries compared to Guinea could make UAV an even more cost-effective solution for EID blood sample transportation.

As time to convey blood samples or results between sites and laboratories remains the main factor of delayed ART initiation among HIV-infected infants in many African contexts(7,8), there is an urgent need for innovative solutions to improve the turnaround times of emergency transportation.

UAV blood sample transportation can improve EID and our simulation suggests that this strategy is cost-effective in the Region of Conakry. UAV blood sample transportation for EID could also be cost-effective in other contexts of high rates of mother to child transmission and long turnaround time for urgent transportation.

## Data Availability

Analyses were conducted using the software R version 4.0.2. (R Core Team. Vienna, Austria). All the study data and code related to the analysis are publicly available on the GitLab repository: https://gitlab.com/air-pop/air-pop_article.git

## Acknowledgements

The study received funding from the *Agence Nationale de Recherches sur le Sida et les hépatites virales/ Maladies Infectieuses Emergentes*—ANRS-MIE (grant number: ANRS 12407).

In addition, we would like to thank the following persons for their valuable contribution. Dr Valériane Leroy and Karen Malateste for providing survival data from IeDEA West Africa. Mohamed Soumah for collecting data on travel time by motorcycle needed to validate *Google Maps* estimations. Drone Volt for providing UAV related data. Prof. Mark Gussy for their useful and meaningful comments on the manuscript.

## Supporting information

Appendix 1: Road and flight distance between health centres and central laboratory (Donka) and number of HIV-positive pregnant women by health facility, 2021.

Appendix 2: Summary figure presenting main parameters and data sources

Appendix 3: Estimating time of arrival at the health centres

Appendix 4: Estimating waiting time before being seen in consultation

Appendix 5: Estimating maximum waiting times among mother present in waiting room

Appendix 6: Estimating the percentage of mother returning to receive the test results for their child

Appendix 7: Estimating the probability of initiating ART after one year in infected children

Appendix 8: Estimating inoperability because of bad weather

Appendix 9: Estimating travel times by monocycles

Appendix 10: Travel time saved by UAV (vs motorcycle) depending on centre distance from the central laboratory

Appendix 11: Survival probability functions for HIV-exposed infants

Appendix 12: Additional inputs related to costs for UAV and motorcycle transportation

## References

1. UNAIDS data 2021. Geneva: Joint United Nations Programme on HIV/AIDS; 2021.

2. Newell ML, Coovadia H, Cortina-Borja M, Rollins N, Gaillard P, Dabis F. Mortality of infected and uninfected infants born to HIV-infected mothers in Africa: a pooled analysis. The Lancet. 2004 Oct 2;364(9441):1236–43.

3. World Health Organization. Guidelines: updated recommendations on HIV prevention, infant diagnosis, antiretroviral initiation and monitoring [Internet]. Geneva: World Health Organization; 2021 [cited 2021 Oct 28]. Available from: https://apps.who.int/iris/handle/10665/340190

4. Bianchi F, Cohn J, Sacks E, Bailey R, Lemaire JF, Machekano R, et al. Evaluation of a routine point-of-care intervention for early infant diagnosis of HIV: an observational study in eight African countries. Lancet HIV. 2019 Jun;6(6):e373–81.

5. Mwenda R, Fong Y, Magombo T, Saka E, Midiani D, Mwase C, et al. Significant Patient Impact Observed Upon Implementation of Point-of-Care Early Infant Diagnosis Technologies in an Observational Study in Malawi. Clin Infect Dis Off Publ Infect Dis Soc Am. 2018 Sep 1;67(5):701–7.

6. Unicef. Key Considerations for Introducing New HIV Point-of-Care Diagnostic Technologies in National Health Systems [Internet]. New York: United National Children’s Fund; 2018 p. 48. Available from: https://www.pedaids.org/wp-content/uploads/2019/02/HIV-Point-of-Care-POC-Diagnostics-Training-Toolkit-.pdf

7. Manumbu S, Smart LR, Mwale A, Mate KS, Downs JA. Shortening Turnaround Times for Newborn HIV Testing in Rural Tanzania: A Report from the Field. PLoS Med. 2015 Nov 3;12(11):e1001897.

8. Phiri NA, Lee HY, Chilenga L, Mtika C, Sinyiza F, Musopole O, et al. Early infant diagnosis and outcomes in HIV-exposed infants at a central and a district hospital, Northern Malawi. Public Health Action. 2017 Jun 21;7(2):83–9.

9. Sibanda EL, Weller IVD, Hakim JG, COWAN FM. The magnitude of loss to follow-up of HIV-exposed infants along the prevention of mother-to-child HIV transmission continuum of care: a systematic review and meta-analysis. AIDS Lond. 2013;27(17):2787–97.

10. Ochieng WO, Ye T, Scheel C, Lor A, Saindon J, Yee SL, et al. Uncrewed aircraft systems versus motorcycles to deliver laboratory samples in west Africa: a comparative economic study. Lancet Glob Health. 2020 Jan 1;8(1):e143–51.

11. Mateen FJ, Leung KHB, Vogel AC, Cissé AF, Chan TCY. A drone delivery network for antiepileptic drugs: a framework and modelling case study in a low-income country. Trans R Soc Trop Med Hyg. 2020 Apr 8;114(4):308–14.

12. Haidari LA, Brown ST, Ferguson M, Bancroft E, Spiker M, Wilcox A, et al. The economic and operational value of using drones to transport vaccines. Vaccine. 2016 Jul 25;34(34):4062–7.

13. Philips N, Blauvelt C, Ziba M, Sherman J, Saka E, Bancroft E, et al. Costs Associated with the Use of Unmanned Aerial Vehicles for Transportation of Laboratory Samples in Malawi. Seattle: VillageReach; 2016.

14. Knoblauch AM, de la Rosa S, Sherman J, Blauvelt C, Matemba C, Maxim L, et al. Bidirectional drones to strengthen healthcare provision: experiences and lessons from Madagascar, Malawi and Senegal. BMJ Glob Health. 2019 Jul 30;4(4):e001541.

15. Nisingizwe MP, Ndishimye P, Swaibu K, Nshimiyimana L, Karame P, Dushimiyimana V, et al. Effect of unmanned aerial vehicle (drone) delivery on blood product delivery time and wastage in Rwanda: a retrospective, cross-sectional study and time series analysis. Lancet Glob Health. 2022 Apr 1;10(4):e564–9.

16. Zailani MA, Azma RZ, Aniza I, Rahana AR, Ismail MS, Shahnaz IS, et al. Drone versus ambulance for blood products transportation: an economic evaluation study. BMC Health Serv Res. 2021 Dec 5;21(1):1308.

17. Bahrainwala L, Knoblauch AM, Andriamiadanarivo A, Diab MM, McKinney J, Small PM, et al. Drones and digital adherence monitoring for community-based tuberculosis control in remote Madagascar: A cost-effectiveness analysis. PLOS ONE. 2020 Jul 7;15(7):e0235572.

18. Amukele TK, Sokoll LJ, Pepper D, Howard DP, Street J. Can Unmanned Aerial Systems (Drones) Be Used for the Routine Transport of Chemistry, Hematology, and Coagulation Laboratory Specimens? PLOS ONE. 2015 Jul 29;10(7):e0134020.

19. Olagunju K. Evaluating traffic congestion in developing countries. A case study of Nigeria. J Chart Inst Logist Transp-Niger. 2015;2(3):23–6.

20. Sidibe L, Ministère du Plan et de la Coopération Internationale, Institut National de la Statistique, Bureau Central de Recensement. Rapport d’analyse des données du RGPH3: Perspectives Demographiques [Internet]. Conakry: Bureau Central de Recensement; 2017 [cited 2021 Oct 18] p. 92. (Recensement Général de la Population et de l’Habitation). Available from: http://www.stat-guinee.org/images/Documents/Publications/INS/rapports_enquetes/RGPH3/RGPH3_mortalite.pdf

21. Sub-Saharan Africa Transport Policy Program. Poverty and urban mobility in Conakry [Internet]. 2004. Report No.: Report 09/04/CK. Available from: https://www.ssatp.org/sites/ssatp/files/publications/PapersNotes/Conakry_en.pdf

22. Wettstein C, Mugglin C, Egger M, Blaser N, Vizcaya LS, Estill J, et al. Missed opportunities to prevent mother-to-child-transmission: systematic review and metaanalysis. AIDS Lond Engl. 2012 Nov 28;26(18):2361–73.

23. Susini A. A Technocritical Review of Drones Crash Risk Probabilistic Consequences and its Societal Acceptance. 7.

24. Iyun V, Technau KG, Vinikoor M, Yotebieng M, Vreeman R, Abuogi L, et al. Variations in the characteristics and outcomes of children living with HIV following universal ART in sub-Saharan Africa (2006–17): a retrospective cohort study. Lancet HIV. 2021 Jun 1;8(6):e353–62.

25. Comité National De Lutte Contre Les IST/VIH/Sida [Guinea]. Analyse des données de survie à 12 mois des personnes sous traitement antirétroviral dans les sites de prise en charge du VIH. Conakry; 2016. Report No.: Rapport 2016.

26. Clinton Health Access Initiative. 2016 antiretroviral (ARV) CHAI reference price list. [Internet]. [cited 2021 Oct 31]. Available from: https://3cdmh310dov3470e6x160esb-wpengine.netdna-ssl.com/wp-content/uploads/2016/11/2016-CHAI-ARV-Reference-Price-List_FINAL.pdf

27. Doherty K, Essajee S, Penazzato M, Holmes C, Resch S, Ciaranello A. Estimating age-based antiretroviral therapy costs for HIV-infected children in resource-limited settings based on World Health Organization weight-based dosing recommendations. BMC Health Serv Res. 2014 Dec;14(1):201.

28. Larson BA, Bii M, Henly-Thomas S, McCoy K, Sawe F, Shaffer D, et al. ART treatment costs and retention in care in Kenya: a cohort study in three rural outpatient clinics. J Int AIDS Soc. 2013;16(1):18026.

29. The World Bank. GDP per capita (current US$), Guinea, Data 2020 [Internet]. The World Bank Website. [cited 2021 Nov 8]. Available from: https://data.worldbank.org/indicator/NY.GDP.PCAP.CD?locations=L3-GN

30. Weinstein MC, Siegel JE, Gold MR, Kamlet MS, Russell LB. Recommendations of the Panel on Cost-effectiveness in Health and Medicine. JAMA. 1996 Oct 16;276(15):1253– 8.

31. McCall B. Sub-Saharan Africa leads the way in medical drones. The Lancet. 2019 Jan 5;393(10166):17–8.

32. Astawesegn FH, Stulz V, Conroy E, Mannan H. Trends and effects of antiretroviral therapy coverage during pregnancy on mother-to-child transmission of HIV in Sub-Saharan Africa. Evidence from panel data analysis. BMC Infect Dis. 2022 Feb 8;22:134.

33. Bhatta M, Dutta N, Nandi S, Dutta S, Saha MK. Mother-to-child HIV transmission and its correlates in India: systematic review and meta-analysis. BMC Pregnancy Childbirth. 2020 Sep 4;20(1):509.

34. Sørensen LY, Jacobsen LT, Hansen JP. Low Cost and Flexible UAV Deployment of Sensors. Sensors [Internet]. 2017 Jan 1 [cited 2022 Feb 2];17(1). Available from: https://europepmc.org/articles/PMC5298727

35. Intelligent Energy showcases drone fuel cell modules in Japan. Fuel Cells Bull. 2019 Aug;2019(8):4–4.

36. Khan KS, Wojdyla D, Say L, Gülmezoglu AM, Van Look PF. WHO analysis of causes of maternal death: a systematic review. Lancet Lond Engl. 2006 Apr 1;367(9516):1066– 74.

37. Bauer J, Moormann D, Strametz R, Groneberg DA. Development of unmanned aerial vehicle (UAV) networks delivering early defibrillation for out-of-hospital cardiac arrests (OHCA) in areas lacking timely access to emergency medical services (EMS) in Germany: a comparative economic study. BMJ Open. 2021 Jan 1;11(1):e043791.

38. Lee J, Wang J, Crandall D, Šabanović S, Fox G. Real-Time, Cloud-Based Object Detection for Unmanned Aerial Vehicles. In: 2017 First IEEE International Conference on Robotic Computing (IRC). 2017. p. 36–43.

