## Supplementary materials for "Improving early infant diagnosis for HIV-exposed infants using unmanned aerial vehicles versus motorcycles for blood sample in Conakry, Guinea: a comparative cost-effectiveness analysis"

**Appendix 1: Road and flight distance between health centres and central laboratory (Donka) and number of HIV-positive pregnant women by health facility, 2021.**

| Health center | Distance (road) from Donka | Distance (flight) from Donka | Pregnant women tested positive for HIV in 2021 |
|---|---|---|---|
| Coleyah | 1.2 | 0.7 | 40 |
| Dixinn | 2.5 | 1.9 | 24 |
| Maciré | 3.3 | 2.5 | 22 |
| Bernard Kouchner | 3.3 | 2.9 | 29 |
| Madina | 3.4 | 2.0 | 87 |
| Koulewondy | 4.7 | 3.9 | 27 |
| Ignace Deen Hospital | 4.8 | 3.9 | 36 |
| Boulbinet | 4.9 | 4.2 | 21 |
| Hafia | 5 | 3.7 | 6 |
| Matam | 5.1 | 4.1 | 55 |
| Fraternité médicale de Guinée | 6.6 | 5.2 | 10 |
| Ratoma | 8.1 | 5.9 | 57 |
| Flamboyant | 8.7 | 7.4 | 61 |
| Gbessia port 1 | 9.7 | 7.6 | 60 |
| Koloma | 11 | 10.0 | 9 |
| Bernay Fotoba | 12.3 | 11.4 | 87 |
| Matoto | 14.7 | 13.8 | 18 |
| Saint Gabriel | 15.4 | 13.6 | 76 |
| Minère | 16.8 | 13.5 | 45 |
| Wanindara | 17 | 15.5 | 43 |
| Lambanyi | 17.1 | 15.1 | 25 |
| Tombolia | 20.3 | 18.6 | 33 |
| Kobaya | 21.6 | 17.4 | 20 |
| Dabompa-DCS Matoto | 23 | 21.3 | 57 |
| Sonfonia | 23.3 | 21.1 | 16 |
| Maneah | 42.4 | 37.1 | 41 |
| Mafoudia | 43.8 | 34.4 | 69 |
| Coyah Hospital | 44.2 | 37.6 | 26 |
| Tanene | 44.6 | 37.1 | 19 |
| Doumbouyah | 45.7 | 37.7 | 30 |
| Fily 2 | 46.4 | 39.1 | 33 |
| Wonkifong | 52 | 36.7 | 9 |
| Khorira | 52.8 | 44.8 | 17 |

#### Appendix 2: Summary figure presenting main parameters and data sources

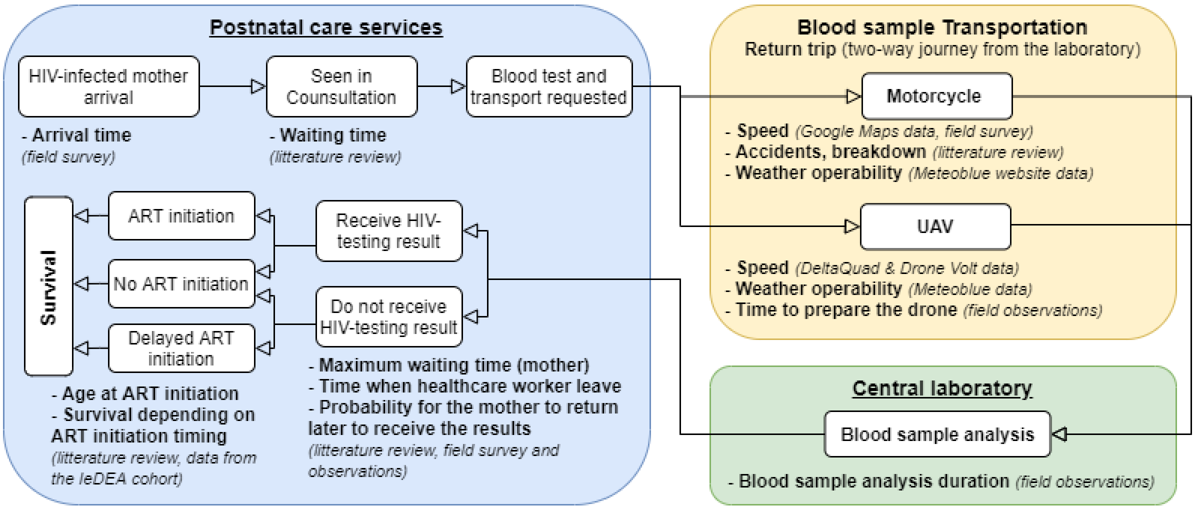

ART: antiretroviral therapy; UAV: unmanned aerial vehicle

### Appendix 3: Estimating time of arrival at the health centres

Because attendance records are poorly detailed in the different sites we included in our study, we collect time of arrival of women visiting two health centres (Figure S3-1). These two data sets show a higher frequency of visits early in the morning, with a steady decrease in the number of visits throughout the day. Women arrive before the opening time of the facilities to benefit from a shorter waiting time. From these observations, we created a fitted distribution that will be used in our model.

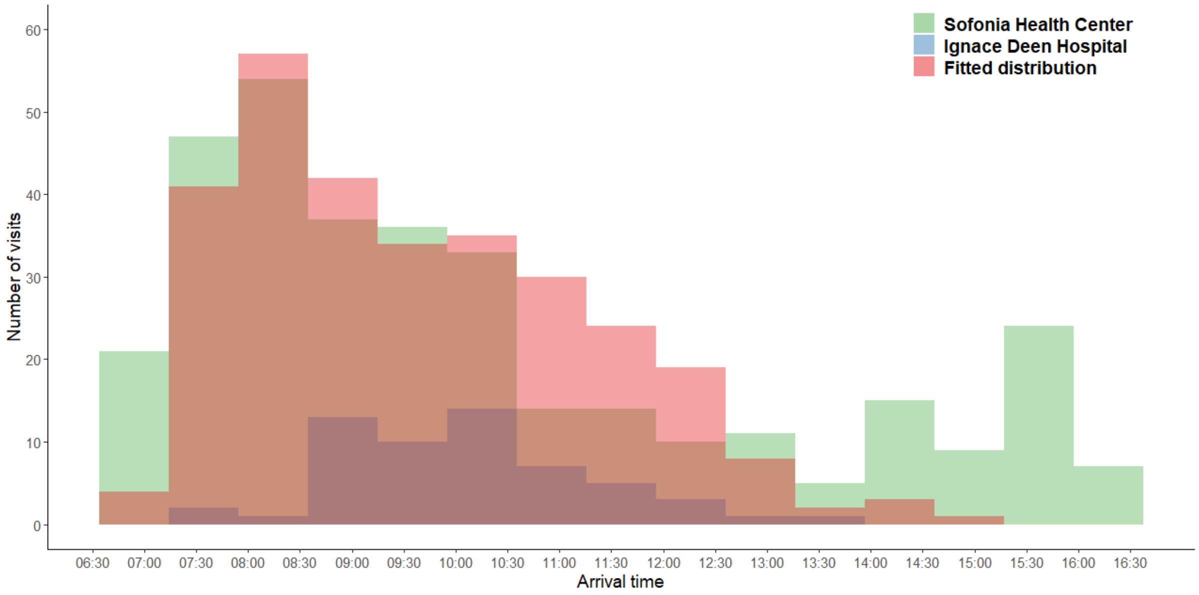

**Figure S3**Error! Reference source not found.Error! Reference source not found.Error! Reference source not found.Error! Reference source not found.**-1: Arrival times at two facilities among women and fitted distribution.**

### Appendix 4: Estimating waiting time before being seen in consultation

The length of time to be seen in consultation was determined from the available literature in the African context.[1–5] The details of the studies included are described in the table below (Table S4-1). From these studies, we calculated a weighted average waiting time of 134 minutes (standard deviation: 66 minutes). Assuming a truncated normal distribution to exclude negative values, the cumulative distribution based on the data obtained from the literature is presented on the right (Figure S4-1).

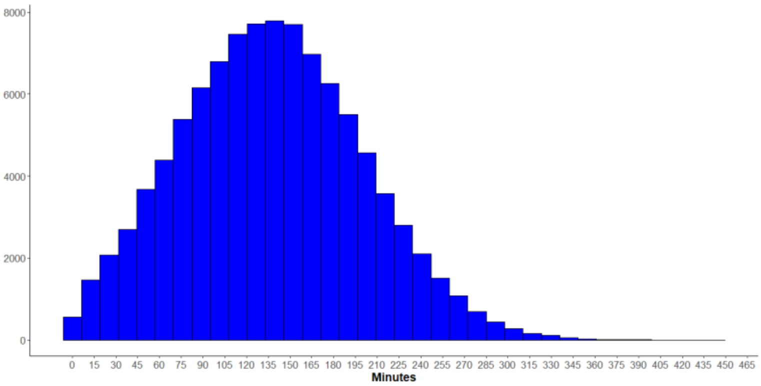

**Figure S4-1 : normal distribution with truncated negative values (mean : 134 ; SD : 66)**

**Table S4-1: List of studies included to estimate waiting time before consultation**

| | | | | **Waiting time before consultation** | | |
|---|---|---|---|---|---|---|
| **Author, year** | **Country** | **Health service** | **size** | Mean | SD | **Comments** |
| Oche MO, 2014 | Nigeria | Outpatient, tertiary hospital | 96 | 161·9 | 109·6 | |
| Steenland M, 2019 | Mozambique | Prenatal services, urban health centre | 505 | 281·6 | 126·3 | We considered only the waiting time before the intervention. |
| | | | 377 | 178·8 | 87·1 | |
| | | | 432 | 175·6 | 93·0 | |
| | | Services prénataux, centre de santé rural | 433 | 128·2 | 86·7 | |
| Umar I, 2011 | Nigeria | Outpatient, tertiary health institute | 384 | 85·0 | 38·75 | As the standard deviation was not available, we recalculated it from the rule of thumb of Ramirez & Cox.[6] |
| Swart AT, 2018 | South Africa | Two primary health centres | 360 | 104·56 | 72·19 | We consider the time on arrival (i.e., registration/reception) with the waiting time before being received in consultation. |
| Do M, 2017 | Kenya | Prenatal services, health centre/hospital | 890 | 71·88 | 2·8075 | Sample sizes were retrieved from the original source document (*DHS Service provision Assessment Kenya and Namibia*) Standard deviation was recalculated from the rule of thumb of Ramirez & Cox.[6] |
| | | Prenatal services, maternity/ dispensary/ clinic | 518 | 75·79 | 6·9725 | |
| | Namibia | Prenatal services, health centre/hospital | 282 | 162·44 | 10·195 | |
| | | Prenatal services, maternity/ dispensary/ clinic | 577 | 127·93 | 5·2425 | |

### Appendix 5: Estimating maximum waiting times among mother present in waiting room

While data on maximum waiting times before a patient decides to leave before being seen are widely available in Northern countries, few published data have been found in Africa. In addition, the available data show some heterogeneity between different settings and types of services.[7–9] To fill this gap, we conducted a short survey among 82 women present in the waiting rooms of pre and postnatal care services of 18 facilities. To consider the variability of responses by facility, we applied a cluster effect to our results.

Two indicators were collected here, the proportion of women who could wait until their child's test result was available (regardless of time) and those who had to leave after a certain time.

Our results showed that 39·0% (32/82, confidence interval CI: [15·3% - 69·0%]) reported being able to wait until the result was available. Of the remainder, we computed the distribution of reported maximum wait times and fitted the associated Beta-PERT distribution (Figure S5-1).

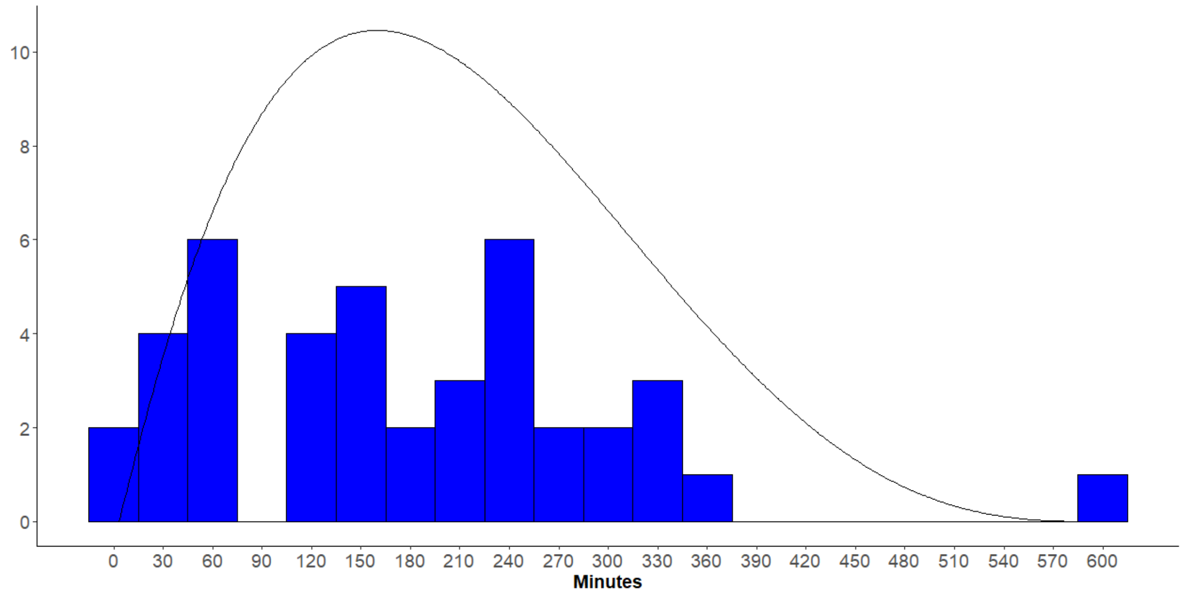

**Figure S5-1: Distribution of the maximum waiting time reported by women seen in the waiting room of 18 health facilities (n= 50).**

#### Appendix 6: Estimating the percentage of mother returning to receive the test results for their child

Assuming that the result would not be returned to the mother of the tested child the same day, we estimated the probability and the time (in days) after which the mother would return to receive the HIV-test result for her child. The mathematical function of the cumulative percentage of women returning to receive the test result as a function of the time between the availability of the test in the centre and the receipt of the test result by the mother was obtained from 5 studies conducted in the African context.[10–14] From these data, we derived a logarithmic function curve using the weighted data for our analysis (Figure S5-1). Under the assumption that no mothers receive their results 1 year after their child's test, this curve gives a median of 33 days [IQR: 5-215] or an average of 59·6 days to receive the results. In comparison, a recent literature review measured an average delay of 44·5 days between the time the test result is available at the health centre and the time mothers retrieve the result.[15]

We make the reasonable assumption that from 12 months onwards, no mothers come to collect the test results. Values above 365 days are thus excluded resulting in 18.0% of mothers not returning for test results in the first year. Another parameter considers the possibility of knowing and initiating a treatment after 365 days (cf. Appendix 7).

In the specific case of an infant leaving before a blood sample was drawn, we assumed that the probabilistic function of the number of days before a mother come back to test her infant for HIV and get the result was equivalent to the probabilistic function described above (Figure S6-1).

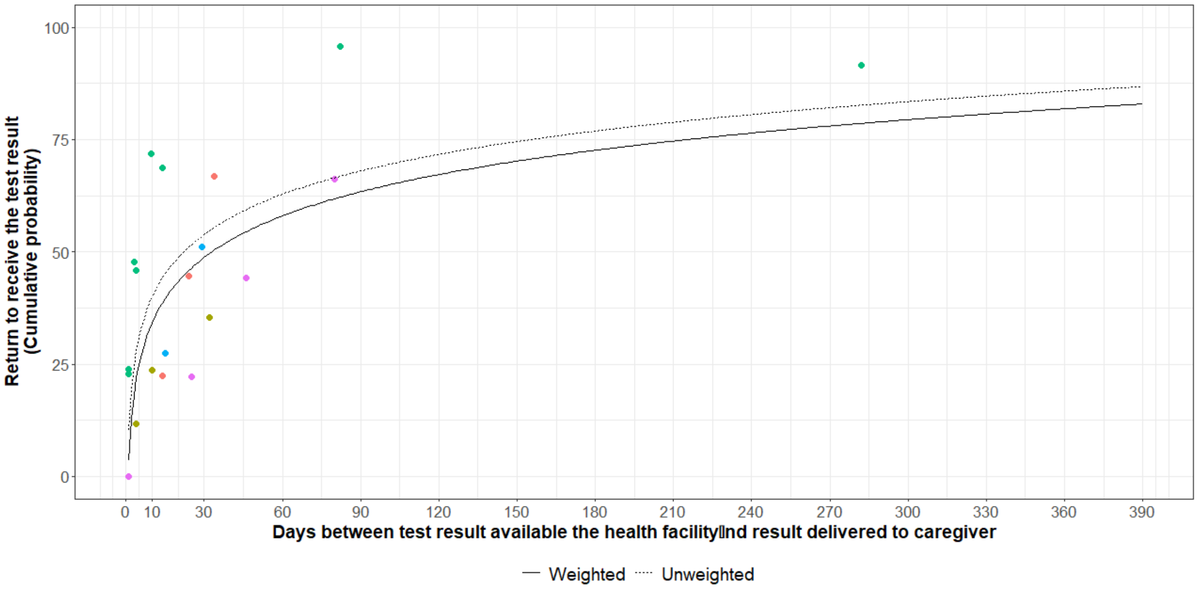

**Figure S6-1: likelihood of receiving test results based on the number of days since the result was available at the health centre.**

Note: each colour represents a specific study.

### Appendix 7: Estimating the probability of initiating ART after one year in infected children

For HIV-infected children less than 1 year old, the probability of initiating ART depends on the probability of receiving the test result (cf. Appendix 6) and the probability of initiating treatment when offered by a health professional. This probability of initiating treatment when offered was set at 92·3% based on the results of a study including 339 care sites in 8 African countries.[16]

For children not initiating ART or those whose test result was not received by the mother within 365 days, we created a function that took into account the probability of initiating treatment after this period. Based on the data from the IeDEA[1] cohort of HIV-infected children in the West Africa, the median age of ART initiation was determined to be 5·6 years [Interquartile range: 2·6-2·9].[17] By setting the minimum value at 1 year and the maximum value at 21·7 years (i.e., the maximum survival age for HIV infection at birth obtained by modelling, cf. Appendix 11), we were able to derive an exponential function for the cumulative probability of initiating ART as a function of time (Figure S7-1).

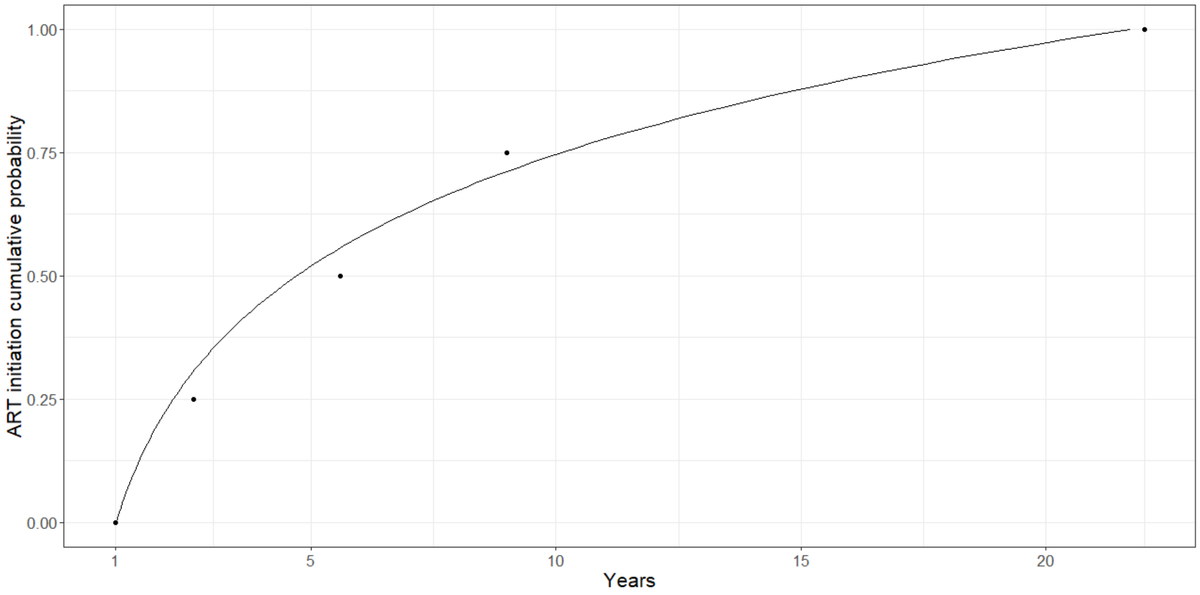

**Figure S7-1: Cumulative probability function curve for initiating antiretroviral therapy (ART) after one year of life.**

[1] For more information: https://www.iedea.org/

### Appendix 8: Estimating inoperability because of bad weather

We collect meteorological data for the city of Conakry during the period 1985-2021 via the website *Meteoblue*[2]. These data included wind speed, gust speed and rainfall volume per day and hour. Over the whole period considered, there was no report of a time window where the wind speed at 80 metres or the wind gusts exceeded 10 km per hour. We therefore neglected the wind strength, as the weather data shows that it has not exceeded the maximum airworthiness limit of our UAV model over the last 20 years.

With respect to precipitation, we estimated the probability of inoperability (i.e., the probability of not being able to use the UAV or motorbike) as a function of the amount of precipitation in certain months and times (Figure S8-1). According to the manufacturer's UAV, the use of the UAV is possible under "light" rainfall - i.e., rainfall less than 2·5 mm per hour according to the American Meteorological Society glossary.[18] Considering the risk of flooding, as well as the danger of driving a motorbike in heavy rain (>7·6 mm/h), we considered this threshold to be the limit of motorcycle operability.

Overall, precipitations are mainly concentrated during the days (Figure S8-1), we then decided to consider only the daytime periods (i.e., between 7am and 7pm), as the transport of blood samples is not supposed to take place during the night. During daytime periods, precipitation have very little influence on motorcycle use (probability of inoperability of 0·2% per trip) and it remains low for UAV (2·6%). This probability of inoperability is nevertheless particularly high in August during the rainy season (1·2% for the motorbike compared to 10·8% for UAV).

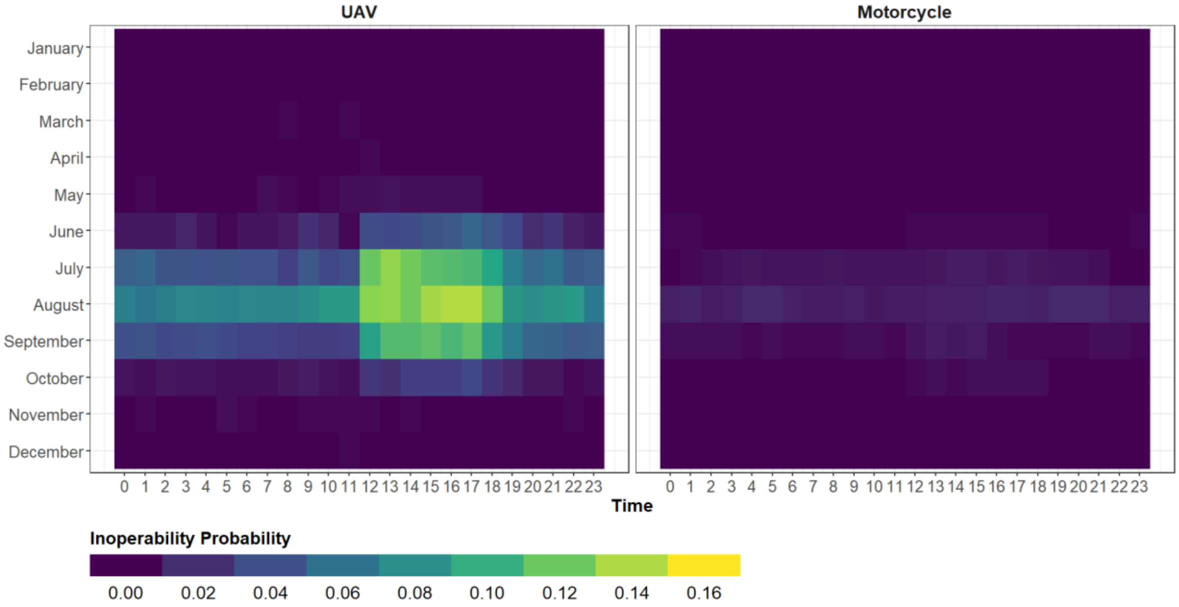

**Figure S8-1: Probability of inoperability due to rainfall when travelling by UAV or motorbike in the city of Conakry, Guinea.**

[2] *Meteoblue* est un site proposant des prévisions météorologiques créé par l'université de Basel en Suisse. Site internet : www.meteoblue.com

### Appendix 9: Estimating travel times by monocycles

Travel times by motorcycles were estimated using Google Maps data. Google Maps travel time estimations depend on departure time and travel direction (Figure S9-1). Thus, we conducted over 353,600 simulations to estimate travel times between the central laboratory and other health centres at various months, day, time slots and directions. Google Maps offer various time of travel estimation (pessimistic, optimistic, or "best-guess" estimates) based on travel time by car. Since motorcycle travel is supposed to be shorter than with car, we considered optimistic and "best-guess" estimates. We validated these estimations by measuring real life travel times by motorcycle –two return trip duration have been collected for each health centres from the central laboratory. Google Maps optimistic estimations were closest to real life estimations and were thus chosen for our analysis (Figure S9-2).

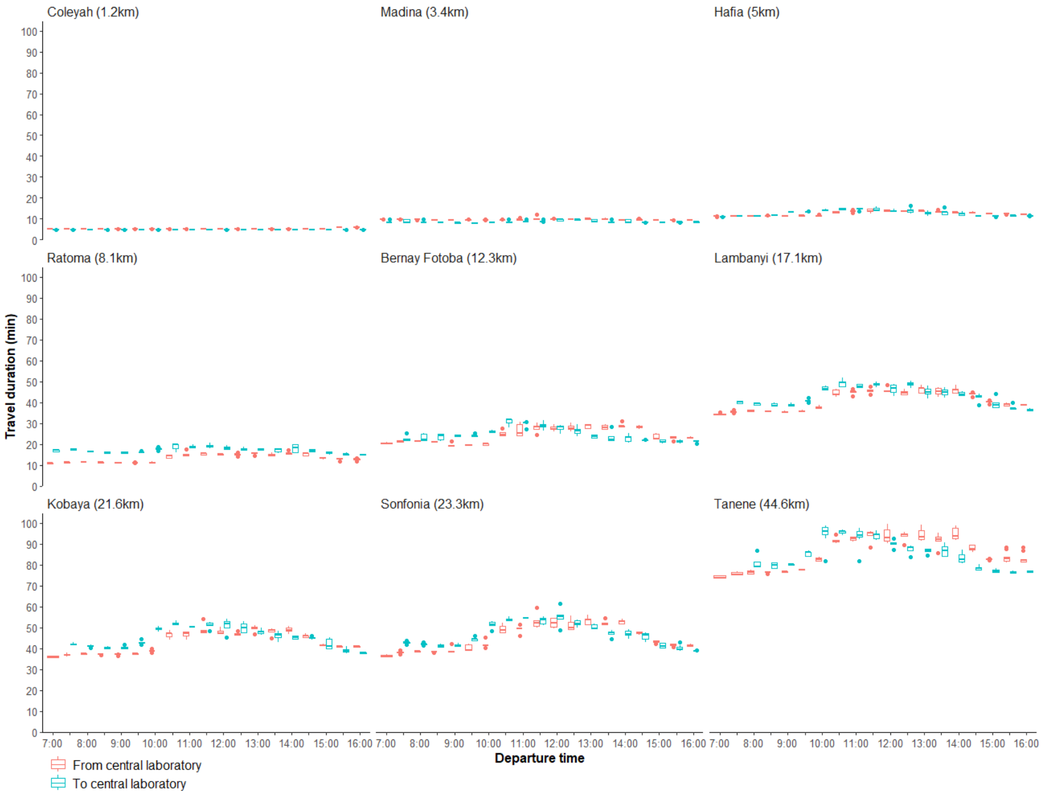

**Figure S9-1: Travel duration from and to the central laboratory by health centres and departure time, Google Maps.**

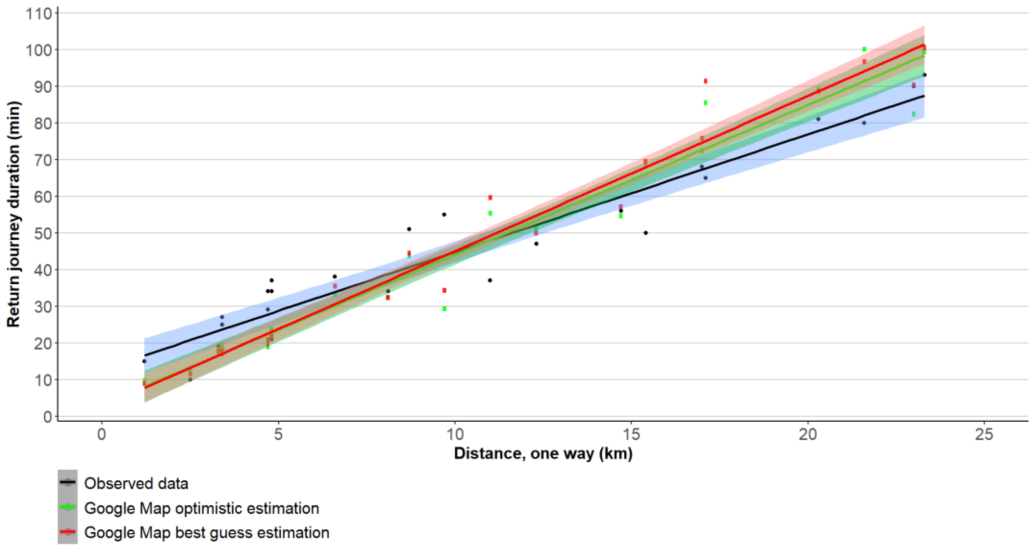

**Figure S9-2: Return journey duration depending on distance, observed and Google Maps data.**

**Appendix 10: Travel time saved by UAV (vs motorcycle) depending on centre distance from the central laboratory**

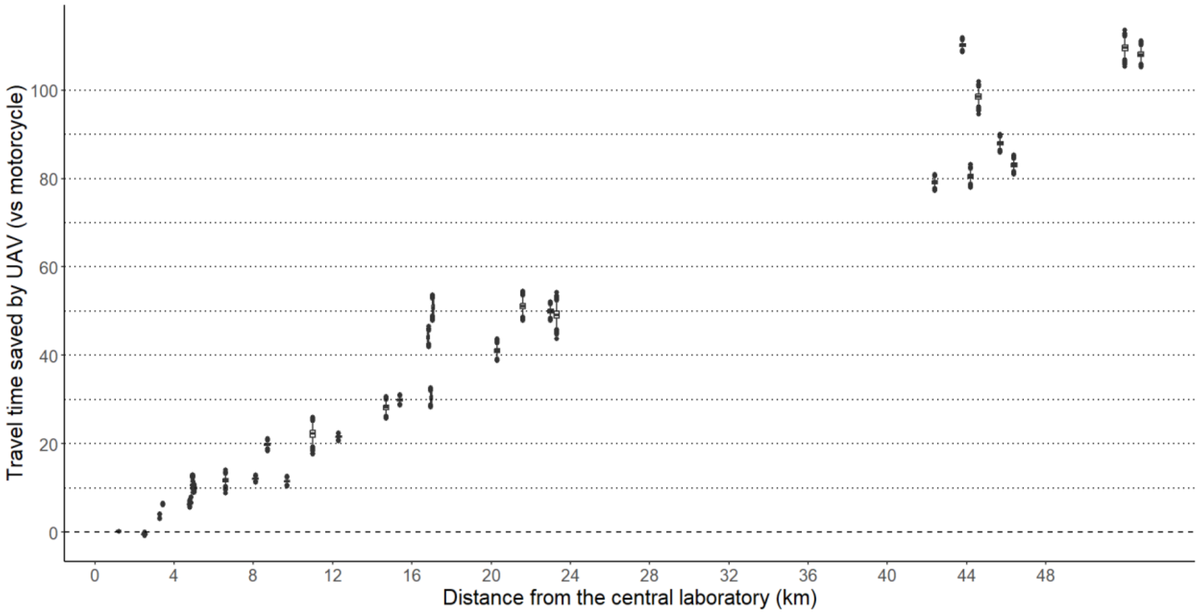

### Appendix 11: Survival probability functions for HIV-exposed infants

We estimated the risk of death curves for the following three groups: (i) HIV-exposed (i.e., HIV-positive mother during pregnancy) but uninfected infants, (ii) Infected infants who did not initiate ART, (iii) Infected infants who initiated ART.

We considered the risk of death from 42 days (6 weeks) of life corresponding to the consultation period recommended by the WHO to perform PCR HIV-testing following exposure to HIV during delivery.[19] The probabilities of death were therefore calculated from 6 weeks.

For all survival curves, we set the maximum survival age at 100 years. This assumption of an age limit of 100 years has little impact on our results, given the negligible percentage of people reaching this age in Northern countries and their extremely low life expectancy at this age.[20] In addition, life expectancy at age 80 in urban Guinea is estimated at 5 years.[21]

#### a) Survival in HIV-exposed but uninfected infants

First, we computed the risk of death curves of HIV-exposed but uninfected infants from the mortality tables of the last general census of the Guinean population conducted in 2014.[21] Since our analysis takes place in Conakry, mortality tables for the urban area were considered. We doubled the mortality risk of dying before age 5 because uninfected infants born to HIV-infected mothers are twice likely to die at 24 months compared to those born to HIV-negative mothers.[22] We assumed that the risk of death after 5 years in uninfected HIV-exposed children was similar to non-exposed children.

As frequently used in survival studies we used a double Weibull distribution[23,24], the double Weibull distribution provides a good functional representation of the human survival curve in many contexts because it allows for a high initial mortality followed by an increasing mortality at later times. The equation for this curve is written as:

$$F(x) = a\left[\frac{\alpha_1}{\beta_1}\left(\frac{x}{\beta_1}\right)^{(\alpha_1-1)} e^{-\left(\frac{x}{\beta_1}\right)^{\alpha_1}}\right] + b\left[\frac{\alpha_2}{\beta_2}\left(\frac{x}{\beta_2}\right)^{(\alpha_2-1)} e^{-\left(\frac{x}{\beta_2}\right)^{\alpha_2}}\right] \quad \alpha, \beta > 0;\ \ a, b \in \mathbb{R} \qquad \textit{Equation (1)}$$

Equation 1 was used to plot the two cumulative risk of death curves for HIV-exposed but uninfected and unexposed infants (Figure S11-1, B). This equation resulted in a life expectancy of 60·3 years for unexposed infants and 53·3 years for HIV-exposed infants at 42 days of life. For comparison, the life expectancy at birth of unexposed children is 59·1 years at birth[21], slightly lower than ours since infant mortality is highest in the first days of life.

#### b) Survival in infected infants without ART initation

The survival function for infected children without ART initiation was built in two stages. Survival between birth and 2·5 years and between 2·5 years and beyond were considered separately due to the lack of data on survival after 2·5 years in infected children without initiation of ART and the particularly high mortality early in the course of infection in neonates compared to adults.

Equation 1 was used to estimate survival from 0 to 2·5 years using survival data from four studies in the sub-Saharan context.[24–27] Survival after 2·5 years was extrapolated from survival data of HIV-infected adults from 4 studies in Africa and one in Asia.[28–32] The survival after 2·5 years was fitted using a linear function (Figure S11-2). We obtained a life expectancy of untreated HIV-infected children of 4·2 years with a maximum life expectancy of 21·7 years, which remains consistent with other studies that have modelled survival in children living with HIV who never initiated an ART.[33–35]

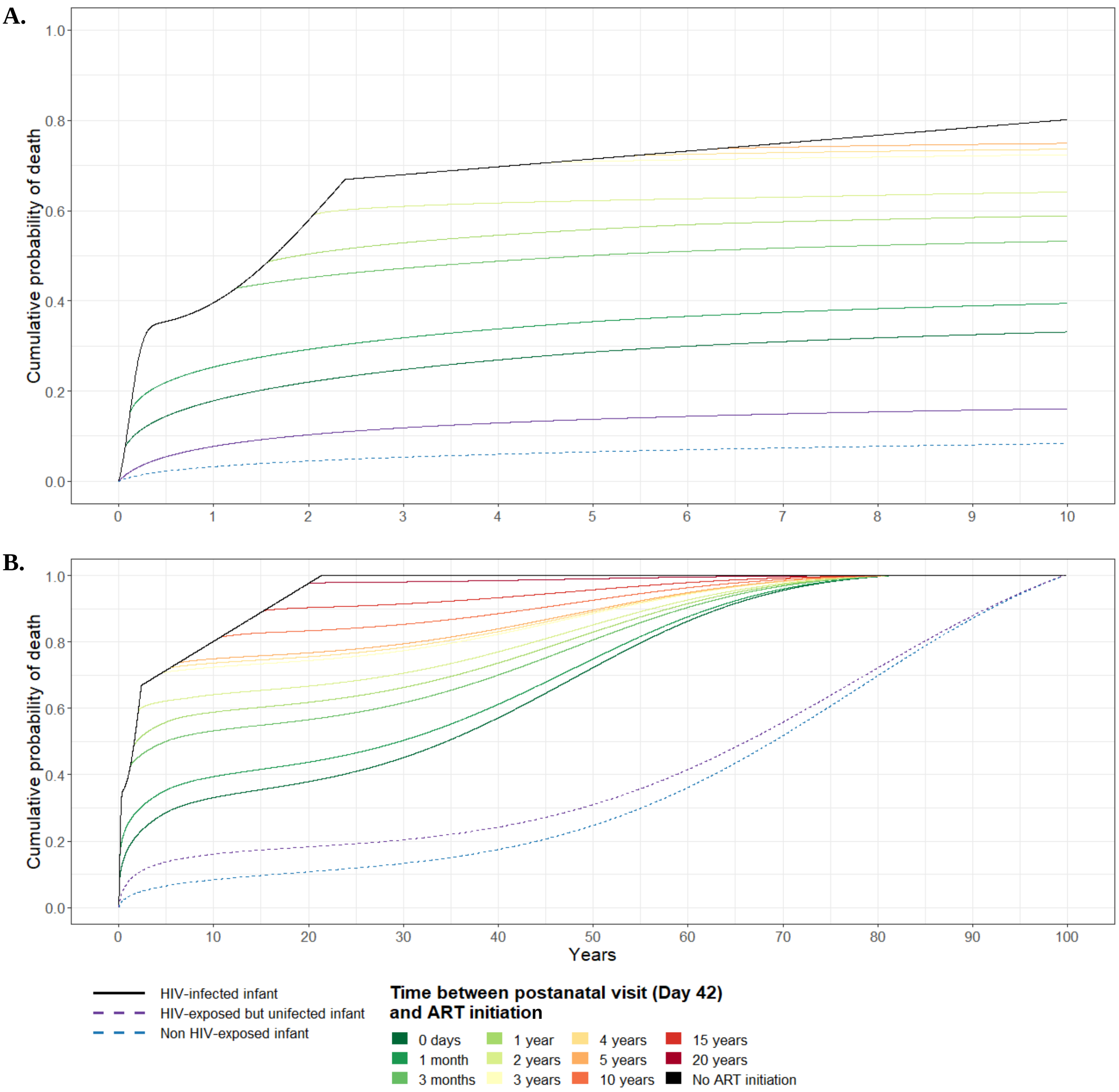

**Figure S11-1: Cumulative probability of death by exposure, HIV status, and age at ART initiation between 42 days of age and 10 years (A) and between 42 days of age and 100 years (B).**

#### c) Survival in HIV-infected infant initiating ART at 6 weeks

Cohort studies of children initiating ART at six weeks rarely exceed 10 years of follow-up, so it is difficult to estimate their life expectancy. Some experts suggests that their life expectancy could be considered to be half that of the national average.[36] Multiplying the probability of death at each age by 4, we obtain a life expectancy at birth of 26·3 years, which is consistent with a study conducted in Zimbabwe (a country with a national life expectancy similar to Guinea), which estimated this life expectancy at 25·5 years.[37] We deduce a life expectancy at 6 weeks of 31·2 years (the life expectancy at 6 weeks is higher than that at birth because of the very high risk of death in the first weeks of life.

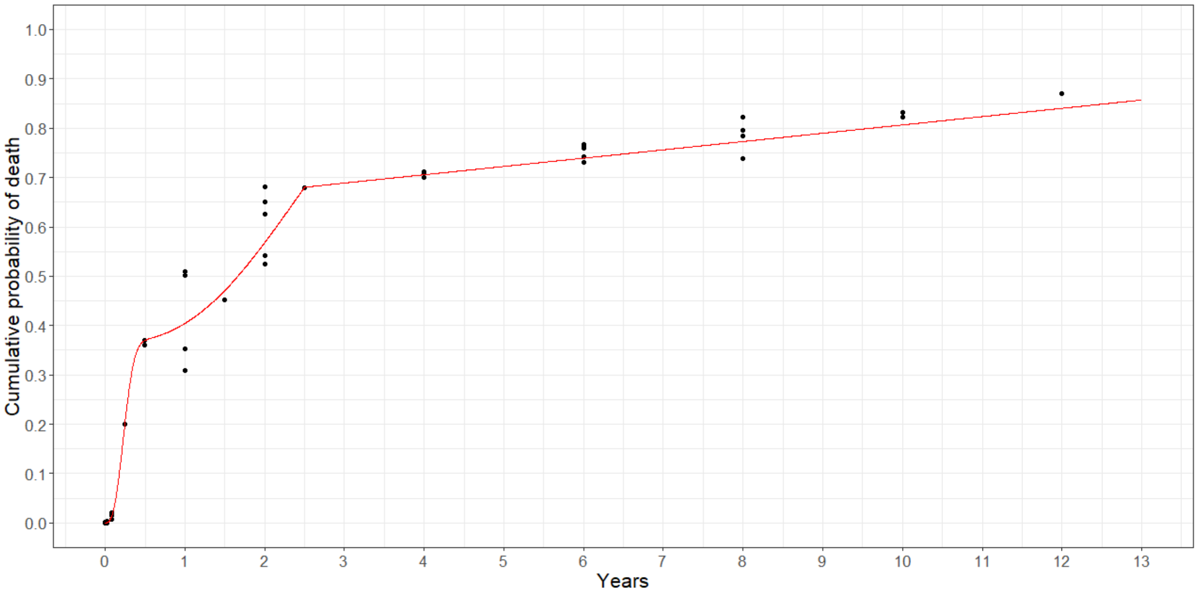

**Figure S11-2: Cumulative probability of death since birth in HIV-infected children not initiating antiretroviral treatment.**

#### d) Survival in HIV-infected children initiating ART after 6 weeks

Data from IeDEA West Africa allowed us to estimate survival curves for children initiating ART between birth and 10 years of age. The risk of death in the year following initiation is different for those who start treatment in the first months of life and those who start at 5 years of age.[38] We created three survival functions depending on the age at ART initiation (≤12 months, 13-60 months, and 61-120 months) as a Weibull distribution (Equation 2).

$$F(x) = \left[\frac{\alpha}{\beta}\left(\frac{x}{\beta}\right)^{(\alpha-1)} e^{-\left(\frac{x}{\beta}\right)^{\alpha}}\right] \quad \alpha,\beta > 0 \qquad Equation\ (2)$$

The cumulative probability of death for a child initiating treatment on day *t* was defined as a function of time spent without ART and time spent on ART depending on age at initiation. Given the paucity of survival data after 60 months of ART initiation in the IeDEA data, we assumed that survival after 60 months on ART followed the same trend as the survival curve for children initiating ART at 6 weeks.

$$F(x) = \begin{cases} x < t, & Equation\ (1)\ for\ infected\ children\ without\ ART \\ t \le x < t + 1825, & Equation\ (2)\ depending\ on\ t\ (\le 365, [366\ - 1825]\ or > 1825) \\ t + 1825\ \le x, & Equation\ (1)\ for\ infected\ children\ initiating\ ART\ at\ D42 \end{cases}$$

To avoid threshold effects at 365 and 1825 days (e.g., higher survival at time *t* for ART initiation at x+1 compared to x) due to the change in the survival function, we assigned any probability $p_t(x)$ greater than $p_{t+1}(x)$ the value of the latter. Figure S11-1 plots the survival function as a function of some value of *t*.

### Appendix 12: Additional inputs related to costs for UAV and motorcycle transportation

| Parameters | Value | Distribution | Sources |
|---|---|---|---|
| **UAV** | | | |
| Rugged Flight case | $1,040 | Point estimate | DeltaQuad |
| Annual maintenance[1] (% of UAV price) | 10% | Point estimate | DeltaQuad |
| Annual insurance cost (% of UAV price) | 10% | | Assumption based on DJI (Shenzhen, Guangdong, China) assurance cost |
| Battery cost (per unit –two needed by UAV) | $577 | Point estimate | DeltaQuad |
| Battery Lifespan (in UAV operating hours) | 1,091 | Point estimate | DeltaQuad. Considering 700 cycles and assuming a 15% maximal autonomy drop. |
| Charging cost for one battery | $0·06 | Point estimate | *Electricité de Guinée*. Estimation based on 340.4 watts/h (14·8 Volts for 23 ampers) and $0·19 per kWh. Three-phase rate for businesses (> 330 kWh/months) |
| UAV pilot/operator training cost (on site course) | $2,891 | Point estimate | DeltaQuad. Include $1,735 on site training in the Netherlands and assumed $1,156 for travel expenses. |
| UAV pilot/operator training cost (online E-course) | $578 | Point estimate | DeltaQuad. |
| Pilot/repair technician salary p.a. | $9,600 | Point estimate | [43] |
| Facility staff training days | 1 | Point estimate | [43] |
| Facility staff training cost per head per day | $58 | Point estimate | [43] |
| Facility staff trained | 66 | Point estimate | Assumption, two staff per facility |
| UAV control console (50km control range) cost | $6358 | Point estimate | DeltaQuad |
| UAV duty fees | $0 | Point estimate | Assumed waived |
| **Motorcycle** | | | |
| Fuel cost (per km) | $0·05 | Point estimate | Estimation based on an operational consumption of 22·1km per litre [43] and $1·03 cost per litre of fuel (local market price) |
| Maintenance (per km) | $0·07 | Point estimate | [43] |
| Annual comprehensive insurance cost | $150 | Point estimate | SUNU assurance, Guinea |
| Vehicle registration document and vignette | $35 | Point estimate | local market price |
| Protective helmet, jacket and gloves | $463 | Point estimate | local market price |
| Healthcare cost for minor accident | $231 | Point estimate | local market price |
| Healthcare cost for minor accident | $1156 | Point estimate | local market price |
| Driver salary p.a. | $4,579 | Point estimate | Solthis. Based on salary grid for dispatch rider in Conakry |
| **Other costs** | | | |
| Temporary replacement cost (% of salary) | 15% | Point estimate | Assumption |
| Cell phone | $19 | Point estimate | local market price |
| Annual cost, mobile plan with 2 hours calls per month | $28 | Point estimate | local market price |

IeDEA; International epidemiology Databases to Evaluate AIDS; PCR: polymerase chain reaction ; SD: standard deviation; Solthis: *Solidarité Thérapeutique et Initiatives pour la Santé*; UAV: unmanned aerial vehicles

[1] Corresponding to a basic tuneup every 12 months (software upgrade, recalibration of sensors, full inspection and test flight, new propellers, motors, speed controllers and battery eliminator circuit) and complete refresh every 24 months (basic turneup with new servos, connectors, board computers, GPS, power module, wiring) as recommended by the manufacturer (https://docs.deltaquad.com/deltaquad-operation-manual/maintenance/scheduled-maintenenance).

### References

1 Oche M, Adamu H. Determinants of patient waiting time in the general outpatient department of a tertiary health institution in north Western Nigeria. *Ann Med Health Sci Res* 2013; **3**: 588–92.

2 Steenland M, Dula J, de Albuquerque A, *et al.* Effects of appointment scheduling on waiting time and utilisation of antenatal care in Mozambique. *BMJ Glob Health* 2019; **4**: e001788.

3 Umar I, Oche M O, Umar A S. Patient waiting time in a tertiary health institution in northern Nigeria. *J Public Health Epidemiol* 2011; **3**: 78–82.

4 Do M, Wang W, Hembling J, Ametepi P. Quality of antenatal care and client satisfaction in Kenya and Namibia. *Int J Qual Health Care J Int Soc Qual Health Care* 2017; **29**: 183–93.

5 Swart A-T, Muller CE, Rabie T. The role of triage to reduce waiting times in primary health care facilities in the North West province of South Africa. *Health SA SA Gesondheid* 2018; **23**: 1097.

6 Ramirez A, Cox C. Improving on the Range Rule of Thumb. ; : 15.

7 Khanna R, Chaudhry MA, Prescott M. Emergency department patients who leave the department without being seen by a doctor. *Eur J Emerg Med* 1999; **6**: 233–5.

8 Liao H-C, Liaw S-J, Hu P, Lee K-T, Chen C-M, Wang F-L. Emergency Department Patients Who Leave without Being Seen by a Doctor: The Experience of a Medical Center in Northern Taiwan. 2002; **25**: 7.

9 Ng Y, Lewena S. Leaving the paediatric emergency department without being seen: Understanding the patient and the risks. *J Paediatr Child Health* 2012; **48**: 10–5.

10 Phiri NA, Lee H-Y, Chilenga L, *et al.* Early infant diagnosis and outcomes in HIV-exposed infants at a central and a district hospital, Northern Malawi. *Public Health Action* 2017; **7**: 83–9.

11 Finocchario-Kessler S, Gautney B, Cheng A, *et al.* Evaluation of the HIV Infant Tracking System (HITSystem) to optimise quality and efficiency of early infant diagnosis: a cluster-randomised trial in Kenya. *Lancet HIV* 2018; **5**: e696–705.

12 Manumbu S, Smart LR, Mwale A, Mate KS, Downs JA. Shortening Turnaround Times for Newborn HIV Testing in Rural Tanzania: A Report from the Field. *PLoS Med* 2015; **12**: e1001897.

13 Sutcliffe CG, Dijk JH van, Hamangaba F, Mayani F, Moss WJ. Turnaround Time for Early Infant HIV Diagnosis in Rural Zambia: A Chart Review. *PLOS ONE* 2014; **9**: e87028.

14 Rollins N, Mzolo S, Moodley T, Esterhuizen T, van Rooyen H. Universal HIV testing of infants at immunization clinics: an acceptable and feasible approach for early infant diagnosis in high HIV prevalence settings. *AIDS Lond Engl* 2009; **23**: 1851–7.

15 Markby J, Boeke C, Sacks J, Wang M, Peter T, Vojnov L. HIV early infant diagnosis testing programs in low- and middle-income countries: a systematic review and meta-analysis. *In preparation*.

16 Bianchi F, Cohn J, Sacks E, *et al.* Evaluation of a routine point-of-care intervention for early infant diagnosis of HIV: an observational study in eight African countries. *Lancet HIV* 2019; **6**: e373–81.

17 Desmonde S, Dicko F, Koueta F, *et al.* Association between age at antiretroviral therapy initiation and 24-month immune response in West-African HIV-infected children. *AIDS* 2014; **28**: 1645–55.

18 American Meteorological Society. Rain. Gloss. Meteorol. https://glossary.ametsoc.org/wiki/Rain (accessed Oct 16, 2021).

19 World Health Organization. Guidelines: updated recommendations on HIV prevention, infant diagnosis, antiretroviral initiation and monitoring. Geneva: World Health Organization, 2021 https://apps.who.int/iris/handle/10665/340190 (accessed Oct 28, 2021).

20 Arias E, Heron M, Xu J. United States Life Tables, 2012. *Natl Vital Stat Rep Cent Dis Control Prev Natl Cent Health Stat Natl Vital Stat Syst* 2016; **65**: 1–65.

21 Diallo MDD, Ministère du Plan et de la Coopération Internationale, Institut National de la Statistique, Bureau Central de Recensement. Rapport d'analyse des données du RGPH3: Mortalité. Conakry: Bureau Central de Recensement, 2017.

22 Arikawa S, Rollins N, Newell M-L, Becquet R. Mortality risk and associated factors in HIV-exposed, uninfected children. *Trop Med Int Health TM IH* 2016; **21**: 720–34.

23 Mudholkar GS, Srivastava DK, Kollia GD. A Generalization of the Weibull Distribution with Application to the Analysis of Survival Data. *J Am Stat Assoc* 1996; **91**: 1575–83.

24 Marston M, Becquet R, Zaba B, *et al.* Net survival of perinatally and postnatally HIV-infected children: a pooled analysis of individual data from sub-Saharan Africa. *Int J Epidemiol* 2011; **40**: 385–96.

25 Marinda E, Humphrey JH, Iliff PJ, *et al.* Child mortality according to maternal and infant HIV status in Zimbabwe. *Pediatr Infect Dis J* 2007; **26**: 519–26.

26 Brahmbhatt H, Kigozi G, Wabwire-Mangen F, *et al.* Mortality in HIV-infected and uninfected children of HIV-infected and uninfected mothers in rural Uganda. *J Acquir Immune Defic Syndr 1999* 2006; **41**: 504–8.

27 Newell M-L, Coovadia H, Cortina-Borja M, Rollins N, Gaillard P, Dabis F. Mortality of infected and uninfected infants born to HIV-infected mothers in Africa: a pooled analysis. *The Lancet* 2004; **364**: 1236–43.

28 Glynn JR, Sonnenberg P, Nelson G, Bester A, Shearer S, Murray J. Survival from HIV-1 seroconversion in Southern Africa: a retrospective cohort study in nearly 2000 gold-miners over 10 years of follow-up. *AIDS* 2007; **21**: 625–32.

29 Urassa M, Boerma JT, Isingo R, *et al.* The impact of HIV/AIDS on mortality and household mobility in rural Tanzania: *AIDS* 2001; **15**: 2017–23.

30 Wawer M, Serwadda D, Gray R, *et al.* Trends in HIV-1 prevalence may not reflect trends in incidence in mature epidemics: data from the Rakai population-based cohort, Uganda. *AIDS* 1997. DOI:10.1097/00002030-199708000-00011.

31 Rangsin R, Chiu J, Khamboonruang C, *et al.* The natural history of HIV-1 infection in young Thai men after seroconversion. *J Acquir Immune Defic Syndr 1999* 2004; **36**: 622–9.

32 Mulder DW, Nunn AJ, Wagner HU, Kamali A, Kengeya-Kayondo JF. HIV-1 incidence and HIV-1-associated mortality in a rural Ugandan population cohort. *AIDS Lond Engl* 1994; **8**: 87–92.

33 Ferrand RA, Corbett EL, Wood R, *et al.* AIDS among older children and adolescents in Southern Africa: projecting the time course and magnitude of the epidemic. *AIDS Lond Engl* 2009; **23**: 2039–46.

34 Stover J, Walker N, Grassly NC, Marston M. Projecting the demographic impact of AIDS and the number of people in need of treatment: updates to the Spectrum projection package. *Sex Transm Infect* 2006; **82 Suppl 3**: iii45-50.

35 Marston M, Zaba B, Salomon JA, Brahmbhatt H, Bagenda D. Estimating the Net Effect of HIV on Child Mortality in African Populations Affected by Generalized HIV Epidemics. *JAIDS J Acquir Immune Defic Syndr* 2005; **38**: 219–27.

36 Aledort JE, Ronald A, Le Blancq SM, *et al.* Reducing the burden of HIV/AIDS in infants: the contribution of improved diagnostics. *Nature* 2006; **444**: 19–28.

37 Frank SC, Cohn J, Dunning L, *et al.* Clinical effect and cost-effectiveness of incorporation of point-of-care assays into early infant HIV diagnosis programmes in Zimbabwe: a modelling study. *Lancet HIV* 2019; **6**: e182–90.

38 Iyun V, Technau K-G, Vinikoor M, *et al.* Variations in the characteristics and outcomes of children living with HIV following universal ART in sub-Saharan Africa (2006–17): a retrospective cohort study. *Lancet HIV* 2021; **8**: e353–62.

39 Comité National De Lutte Contre Les IST/VIH/Sida [Guinea]. Analyse des données de survie à 12 mois des personnes sous traitement antirétroviral dans les sites de prise en charge du VIH. Conakry, 2016.

40 Clinton Health Access Initiative. 2016 antiretroviral (ARV) CHAI reference price list. https://3cdmh310dov3470e6x160esb-wpengine.netdna-ssl.com/wp-content/uploads/2016/11/2016-CHAI-ARV-Reference-Price-List_FINAL.pdf (accessed Oct 31, 2021).

41 Doherty K, Essajee S, Penazzato M, Holmes C, Resch S, Ciaranello A. Estimating age-based antiretroviral therapy costs for HIV-infected children in resource-limited settings based on World Health Organization weight-based dosing recommendations. *BMC Health Serv Res* 2014; **14**: 201.

42 Larson BA, Bii M, Henly-Thomas S, *et al.* ART treatment costs and retention in care in Kenya: a cohort study in three rural outpatient clinics. *J Int AIDS Soc* 2013; **16**: 18026.

43 Ochieng WO, Ye T, Scheel C, *et al.* Uncrewed aircraft systems versus motorcycles to deliver laboratory samples in west Africa: a comparative economic study. *Lancet Glob Health* 2020; **8**: e143–51.
